# AWS Trainium vs NVIDIA CUDA for Medical Image Classification: A Comprehensive Benchmark on ChestX-ray14

**DOI:** 10.64898/2025.12.23.25342933

**Authors:** George Fisher

## Abstract

We present a rigorous benchmark comparing AWS Trainium (trn1 instances) and NVIDIA CUDA (g5 instances with A10G GPUs) for training convolutional neural networks on medical image classification. Using the NIH ChestX-ray14 dataset with 112,120 chest radiographs and 14 thoracic disease labels, we evaluate ResNet-50 and ConvNeXt architectures across both platforms. Our key findings are threefold: (1) Trainium achieves virtually identical accuracy to CUDA for compatible architectures (ConvNeXt-Pico: F1=0.8007 vs 0.8027, Δ=0.25%), (2) modern CNN architectures using depthwise convolutions and LayerNorm (ConvNeXt-Tiny and larger) fail to compile or load on Trainium due to hardware constraints, and (3) Trainium is 3–5 × more expensive than CUDA for CNN training even with correct instance sizing. We document the substantial porting effort required, including four critical XLA-specific code modifications, and provide guidance for practitioners considering Trainium for computer vision workloads.

## 1 Introduction

The rapid growth of deep learning in medical imaging has driven demand for cost-effective training infrastructure. AWS Trainium, announced as a purpose-built machine learning accelerator, promises competitive performance at lower cost than traditional GPU instances [1]. However, most published benchmarks focus on large language models (LLMs) and transformer architectures, leaving uncertainty about Trainium’s suitability for convolutional neural networks (CNNs) in medical imaging applications.

This study provides a rigorous empirical comparison of AWS Trainium and NVIDIA A10G GPUs for training CNNs on the ChestX-ray14 dataset [2], a standard benchmark for thoracic disease classification. We evaluate both classic (ResNet-50) and modern (ConvNeXt) architectures, measuring accuracy, training throughput, compilation overhead, and total cost of ownership.

This study extends our prior work on ChestX-ray14 classification [3], which achieved state-ofthe-art results (F1=0.821, ROC-AUC=0.940) using ConvNeXt ensembles on CUDA. We adapt that training and evaluation pipeline to benchmark AWS Trainium, enabling direct comparison under identical methodology.

Our contributions include:

- We present the first detailed benchmark documenting architecture compatibility, porting requirements, and cost comparison for 2D classification tasks. (AWS reports successful Trainium deployments for 3D medical imaging at MIT [4])
- Documentation of architecture compatibility limits (ConvNeXt-Pico is the largest working ConvNeXt variant)
- Identification of four critical XLA code modifications required for correct training
- Comprehensive cost analysis revealing Trainium is 3–5× more expensive than CUDA for CNNs
- Evidence that Trainium achieves equivalent accuracy when models successfully compile

### 2 Background

### 2.1 AWS Trainium Architecture

AWS Trainium is a custom machine learning accelerator designed by Amazon’s Annapurna Labs [5]. Each Trainium chip contains two NeuronCores, with the trn1.2xlarge instance providing 2 NeuronCores (32GB HBM) and trn1.32xlarge providing 32 NeuronCores (512GB HBM) [6].

Unlike NVIDIA GPUs that execute PyTorch operations eagerly, Trainium uses XLA (Accelerated Linear Algebra) compilation through the torch_xla library [7]. This requires tracing the computation graph and compiling it to Neuron Executable File Format (NEFF) before execution—a process that can take minutes to hours for complex models.

### 2.2 ChestX-ray14 Dataset

The NIH ChestX-ray14 dataset contains 112,120 frontal-view chest radiographs from 30,805 unique patients, with 14 thoracic disease labels extracted from radiology reports using natural language processing [2]. The multi-label classification task is challenging due to label noise, class imbalance (prevalence ranges from 0.2% for Hernia to 19.4% for Infiltration), and the subtle visual features distinguishing pathologies.

### 2.3 Model Architectures

We evaluate two CNN families:

#### ResNet-50 [8]

A classic architecture using standard 3× 3 convolutions, batch normalization, and ReLU activations. With 23.5M parameters, it represents well-established CNN design principles.

#### ConvNeXt [9]

A modernized CNN incorporating design elements from Vision Transformers, including 7×7 depthwise convolutions, LayerNorm, and GELU activations. We test variants from ConvNeXt-Atto (3.7M parameters) to ConvNeXt-Tiny (28M parameters) [10].

## 3 Methods

We use the training and evaluation framework from [3], including patient-level data splits and comprehensive per-disease metrics. This ensures methodological consistency and enables direct comparison with our previously published CUDA results.

### 3.1 Experimental Setup

All experiments used identical hyperparameters across platforms:

- **Resolution:** 128×128 pixels
- **Batch size:** 32
- **Optimizer:** AdamW (learning rate 0.0001, weight decay 10^−4^)
- **Loss:** Binary cross-entropy with logits
- **Early stopping:** Patience of 21 epochs on validation F1, allowing the learning rate scheduler (patience=5) multiple opportunities to reduce LR before termination
- **Data splits:** Patient-level stratified (70% train, 15% validation, 15% test)
- **Random Seed**: set_training_optimizations(seed=42)
- **Classification threshold**: 0.1

### 3.2 Hardware Configuration

### 3.3 Software Environment

#### CUDA platform

PyTorch 2.3.0, CUDA 12.1, FP16 automatic mixed precision with fused AdamW optimizer.

#### Trainium platform

PyTorch 2.8.0, torch_xla 2.8.1, Neuron SDK 2.21, FP32 precision (BF16 showed numerical instability in early experiments).

### 3.4 Naming Convention

Results are organized using the naming convention:

`{platform}_{model}_{resolution}_{precision}_{instance}`

For example, `trainium_convnext_pico_128_fp32_trn1.32xlarge` indicates ConvNeXt-Pico trained at 128×128 resolution in FP32 on a trn1.32xlarge instance.

### 3.5 XLA-Specific Code Modifications

Porting PyTorch code from CUDA to Trainium required four critical modifications, discovered through systematic debugging over 16 sessions (~50 hours):

#### 3.5.1 Optimizer Step

Standard `optimizer.step()` must be replaced with XLA-specific synchronization:

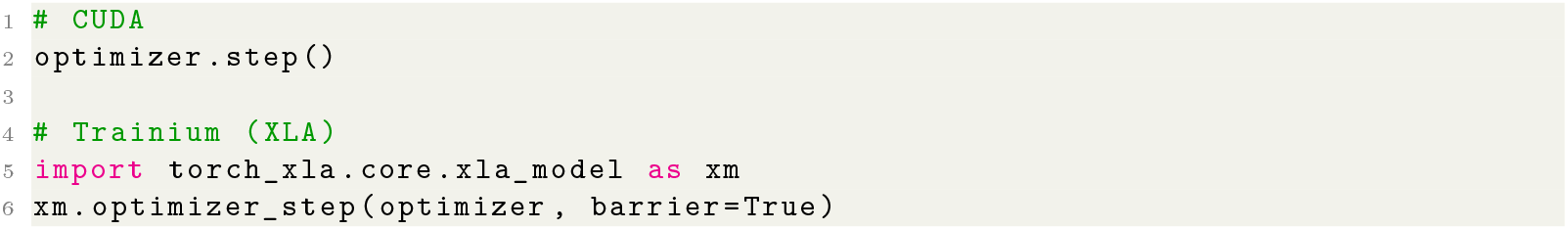

#### 3.5.2 Data Loading

Standard PyTorch DataLoaders must be wrapped for proper device placement:

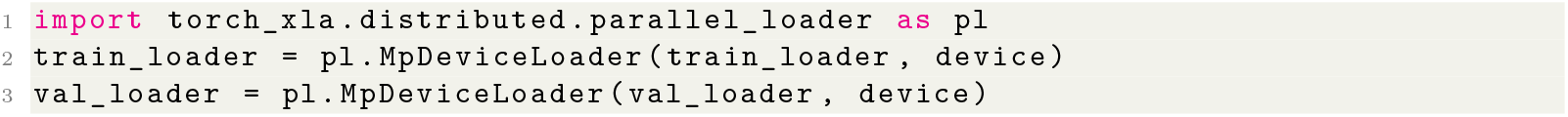

#### 3.5.3 Uniform Batch Sizes

Variable batch sizes (e.g., final incomplete batch) trigger recompilation:

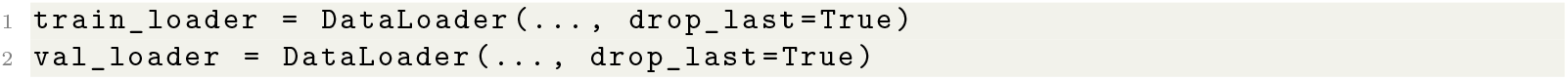

#### 3.5.4 Validation Loop Graph Explosion

The most subtle issue: accumulating predictions in a Python list causes XLA to trace an ever-growing graph, resulting in 30+ minute compilation times. The solution is to force execution and CPU transfer after each batch:

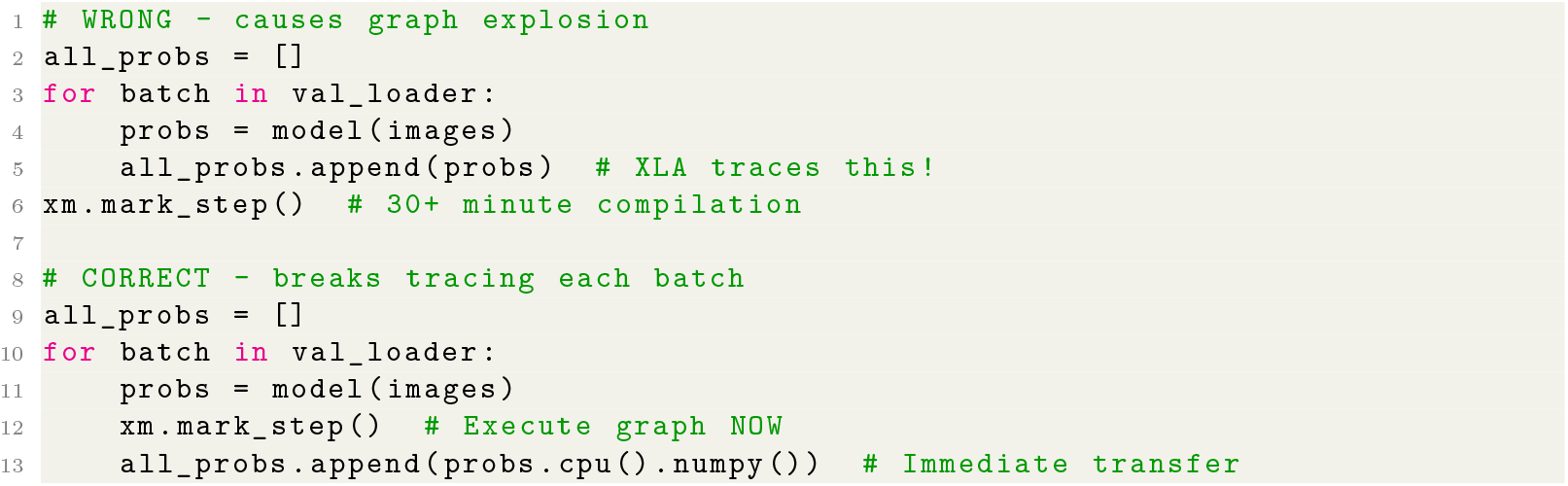

## 4 Results

### 4.1 Architecture Compatibility

Not all architectures successfully compile and run on Trainium. Table 2 summarizes our findings.

**Table 1.** Instance specifications and costs.

| Instance | Accelerator | Memory | Cost/hr | Platform |
| --- | --- | --- | --- | --- |
| g5.2xlarge | 1 NVIDIA A10G | 24GB GDDR6 | \$1.21 | CUDA |
| trn1.2xlarge | 2 NeuronCores | 32GB HBM | \$1.34 | Trainium |
| trn1.32xlarge | 32 NeuronCores | 512GB HBM | \$21.50 | Trainium |

**Table 2.** ConvNeXt variant compatibility with Trainium.

| Variant | Params | Trainium | CUDA | Failure Mode |
| --- | --- | --- | --- | --- |
| ConvNeXt-Atto | 3.7M | ✓ | ✓ | — |
| ConvNeXt-Femto | 5.2M | ✓ | ✓ | — |
| ConvNeXt-Pico | 9.1M | ✓ | ✓ | — |
| ConvNeXt-Nano | 15M | × | ✓ | Scratchpad buffer overflow |
| ConvNeXt-Tiny | 28M | × | ✓ | SB overflow + compiler bugs |

The failure mode for larger ConvNeXt variants is a hardware constraint: the Trainium Scratchpad Buffer (SB) has a ~ 24MB per-instruction limit. ConvNeXt’s architectural components— 7×7 depthwise convolutions, LayerNorm backward pass, and GELU—require more intermediate tensor storage than this limit allows.

#### Key finding

ConvNeXt-Pico (9.1M parameters) is the largest ConvNeXt variant that successfully trains on Trainium.

### 4.2 Accuracy Comparison

For architectures that compile successfully, Trainium achieves virtually identical accuracy to CUDA.

**Table 3.** ConvNeXt-Pico accuracy comparison (FP32, 128×128)

| Metric | CUDA | Trainium | $\Delta$ |
| --- | --- | --- | --- |
| Test F1 | 0.8027 | 0.8007 | −0.25% |
| Test ROC-AUC | 0.8984 | 0.8914 | −0.78% |
| Test PR-AUC | 0.7835 | 0.7569 | −3.4% |
| Test Sensitivity | 0.7190 | 0.7187 | −0.04% |
| Test Specificity | 0.9945 | 0.9940 | −0.05% |
| Test Precision | 0.9108 | 0.9060 | −0.53% |
| Best Validation F1 | 0.7769 | 0.7791 | +0.28% |

The 0.25% F1 difference is within random variation and not statistically significant. This demonstrates that **Trainium produces equivalent model quality when architectures are compatible**.

**Table 4.**
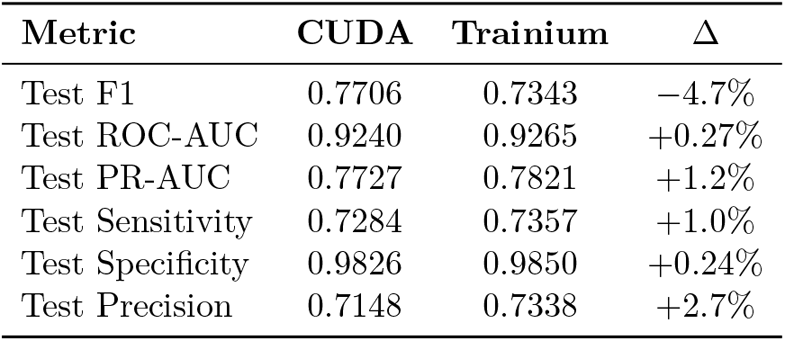
ResNet-50 accuracy comparison (FP32, 128×128)

Interestingly, Trainium achieves slightly *higher* ROC-AUC, PR-AUC, sensitivity, specificity, and precision than CUDA despite lower F1. Since the same fixed threshold (0.1) was used for both platforms, this apparent paradox likely reflects random variation from training dynamics (weight initialization, batch ordering) rather than a fundamental platform difference. The similar ROC-AUC values (0.924 vs 0.927) confirm equivalent discriminative ability; the F1 gap warrants investigation with multiple seeds.

While only a single seed was used due to compilation cost constraints on Trainium, the near-identical ROC-AUC values across platforms and consistency with prior CUDA-only multi-seed results [3] suggest that observed differences fall within expected stochastic variation.

### 4.3 Training Performance

Trainium is consistently slower per epoch, though the gap varies by architecture.

**Table 5.** Per-epoch training time comparison (steady-state, post-compilation)

| Model | CUDA | Trainium | Ratio |
| --- | --- | --- | --- |
| ResNet-50 | 2.1 min | 10.3 min | 4.9× slower |
| ConvNeXt-Pico | 2.5 min | 5.7 min | 2.3× slower |

### 4.4 Compilation Overhead

The compilation overhead is amortized over training epochs but represents a significant cost for failed compilations. Our ConvNeXt-Tiny compilation attempt consumed 14 hours of trn1.32xlarge time (~$300) before failing at model load.

**Table 6.** First-epoch compilation time.

| Model | CUDA | Trainium |
| --- | --- | --- |
| ResNet-50 | 2.3 sec | 5 min |
| ConvNeXt-Pico | <1 min | 15 min |
| ConvNeXt-Tiny | <1 min | 14 hours (then fails) |

### 4.5 Instance Sizing Discovery

A critical finding: **trn1.2xlarge and trn1.32xlarge produce identical per-epoch training times for single-model training**.

**Table 7.** Instance size comparison (Trainium)

| Model | trn1.32xlarge | trn1.2xlarge | Speed Diff |
| --- | --- | --- | --- |
| ResNet-50 | 10.5 min/ep | 10.3 min/ep | None |
| ConvNeXt-Pico | 5.7 min/ep | 5.7 min/ep | None |

The extra 30 NeuronCores on trn1.32xlarge provide zero benefit for single-model training— they are designed for distributed training across multiple model replicas. This discovery reduced our projected Trainium costs by 16×.

### 4.6 Cost Analysis

**Table 8.** Total training cost comparison (ConvNeXt-Pico, 200 epochs)

| Platform | Instance | Time | Cost |
| --- | --- | --- | --- |
| CUDA | g5.2xlarge | 4 hrs | \$5 |
| Trainium (optimal) | trn1.2xlarge | 20 hrs | \$27 |
| Trainium (actual) | trn1.32xlarge | 20 hrs | \$430 |

#### With correct instance sizing, Trainium is 5.4× more expensive than CUDA for CNN training

Our actual costs were 86× higher due to initially using trn1.32xlarge unnecessarily.

With hindsight, optimal execution using trn1.2xlarge from the start the trainium portion would have cost ~$100 instead of $2,324.

### 4.7 Per-Disease Performance

Figure 2 shows per-disease classification performance for the ConvNeXt-Pico model trained on Trainium. Performance varies substantially across diseases (F1 range: 0.749–0.909), influenced by factors including visual distinctiveness and label quality. Notably, Hernia achieves the highest F1 (0.909) despite very low prevalence (0.2%), while Infiltration—the most common finding (19.4%)—shows lower F1 (0.749), likely due to its subtle and diffuse radiographic appearance.

**Figure 1.**
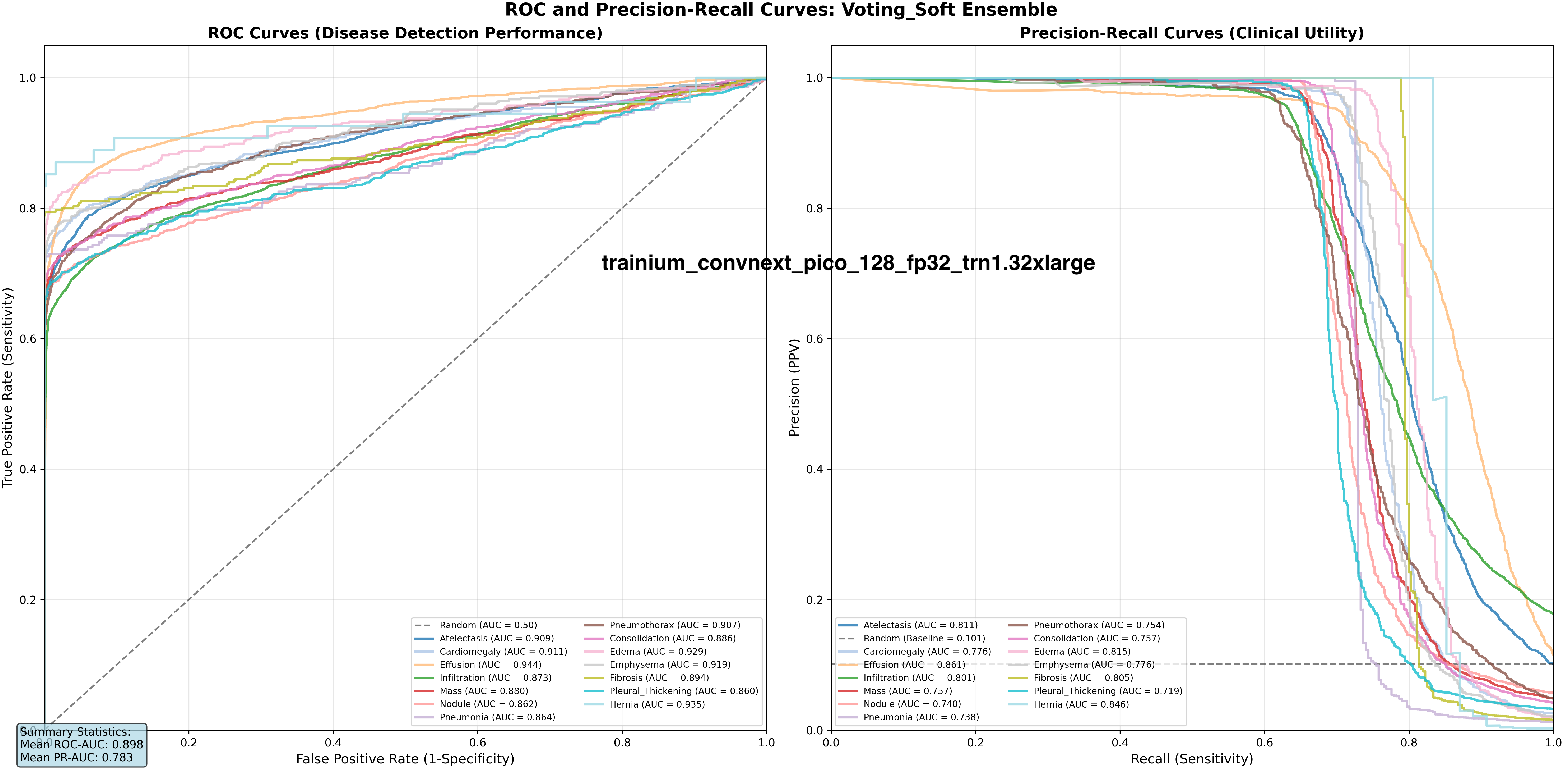
ROC and Precision-Recall curves for ConvNeXt-Pico trained on Trainium. Left: ROC curves show strong discriminative performance across all 14 diseases (mean ROC-AUC = 0.898). Right: Precision-Recall curves demonstrate clinical utility (mean PR-AUC = 0.783), with performance varying substantially across diseases due to factors including visual distinctiveness and label quality.

**Figure 2.**
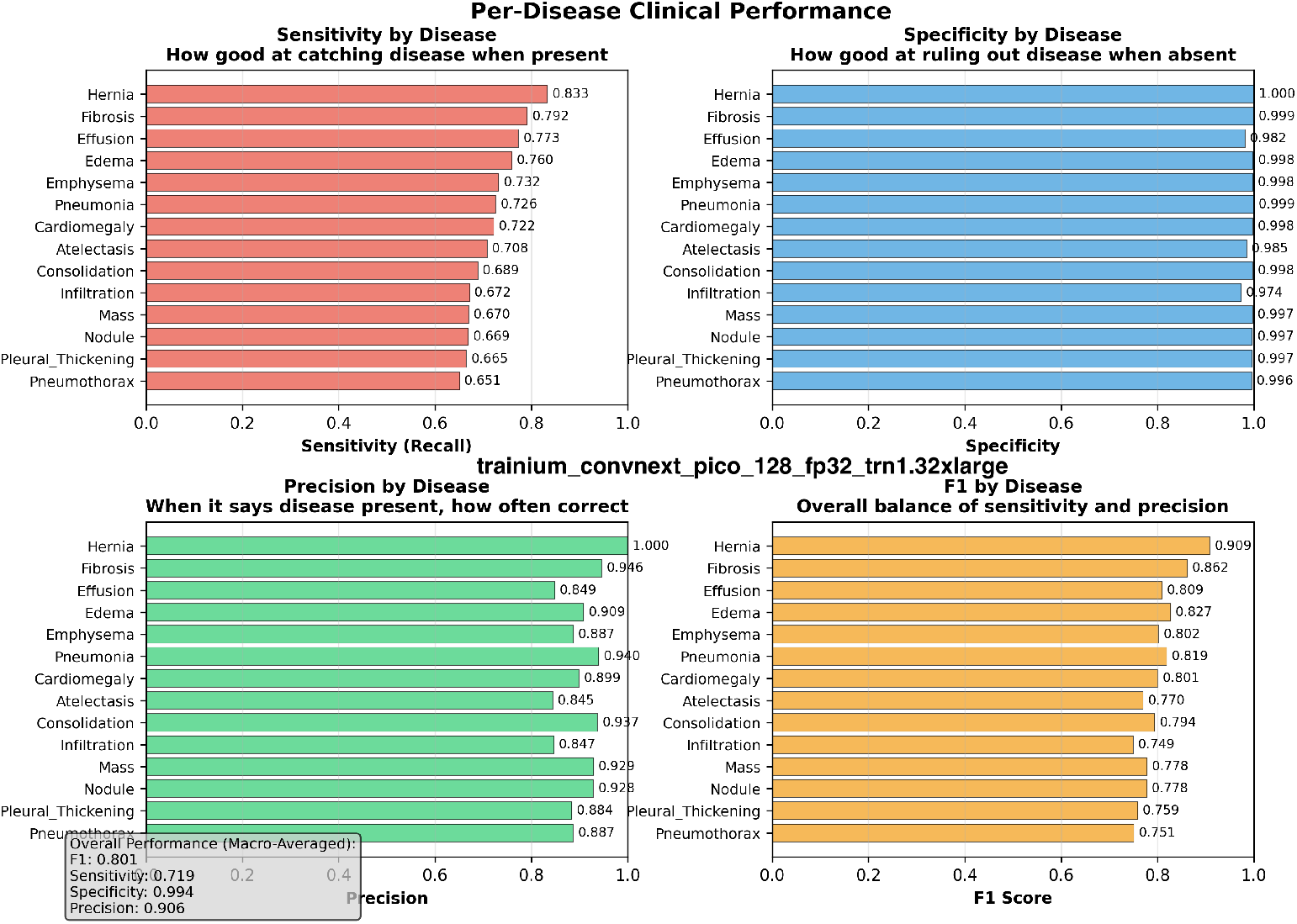
Per-disease clinical performance metrics for ConvNeXt-Pico trained on Trainium

### 4.8 Training Dynamics

Figure 3 shows the training progression for ConvNeXt-Pico on Trainium. The model achieves best validation F1 of 0.7791 at epoch 190, with training loss approaching zero while validation loss stabilizes around 0.20—indicating appropriate regularization without severe overfitting.

**Figure 3.**
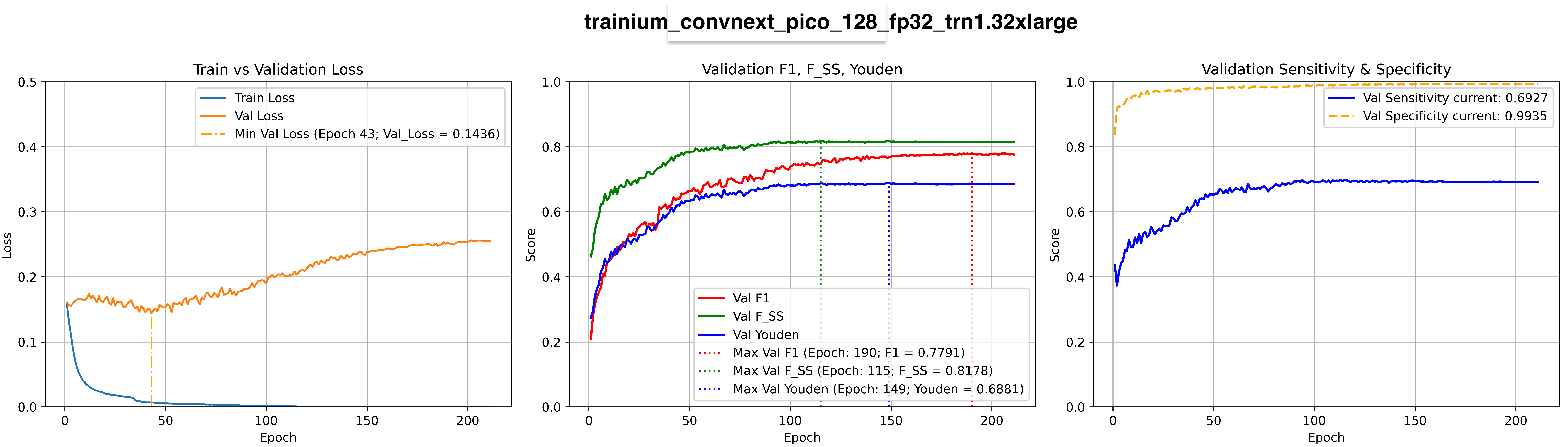
Training curves for ConvNeXt-Pico on Trainium showing loss, validation metrics, and sensitivity/specificity over 211 epochs

Quantitative analysis of training stability reveals Trainium exhibits higher volatility than CUDA. Using Excess Total Variation (ExcessTV)—a measure of training curve “wiggliness” beyond net progress (see Appendix C)—Trainium shows 1.56× higher volatility than CUDA (Trainium ExcessTV = 0.72, CUDA = 0.46). The post-warmup phase (epochs 50+) shows an even larger gap, with volatility ratios of 5.47× vs 3.47×. This increased oscillation may stem from differences in XLA graph execution patterns or numerical accumulation behavior. Practitioners should consider higher early stopping patience on Trainium to avoid premature termination during transient F1 dips.

## 5 Discussion

### 5.1 Architecture Compatibility

Our results reveal a clear pattern: Trainium handles “classic” CNN components (standard convolutions, BatchNorm, ReLU) well, but struggles with modern architectural innovations. The specific problematic components are:

1. **7**×**7 depthwise convolutions:** Create many separate DMA operations that exhaust the descriptor ring buffer
2. **LayerNorm backward pass:** Requires more intermediate tensor storage than BatchNorm
3. **GELU activation:** Compiles to multiple intermediate operations

This suggests Trainium’s architecture was optimized for transformer attention patterns rather than modern CNN designs.

### 5.2 Cost-Effectiveness

Despite AWS marketing Trainium as cost-effective, our analysis shows it is **3–5**× **more expensive than CUDA for CNN training**, even with optimal instance sizing. The cost disadvantage stems from:

1. Slower per-epoch training (2.3–4.9×)
2. Higher hourly cost for comparable performance (trn1.2xlarge vs g5.2xlarge)
3. Compilation overhead (not amortized for short experiments)
4. Risk of failed compilations consuming resources

Trainium may be cost-effective for LLM training where it was specifically optimized, but this does not transfer to CNN workloads.

### 5.3 Porting Effort

The porting effort was substantial: 12 days, 16 sessions, ~50 hours of engineering time, plus $2,750 in compute and AI assistance costs. The four critical XLA modifications we identified are not well-documented, particularly the validation loop graph explosion issue that required AI assistance (Claude.ai) to diagnose.

For production deployments, organizations should budget significant engineering time for Trainium porting, beyond what would be required for multi-GPU CUDA scaling.

### 5.4 Recommendations

Based on our findings:

**Use Trainium for:**

- LLM training (Trainium’s target market)
- Architectures using standard operations (conv2d, BatchNorm, ReLU)
- Distributed training at scale where NeuronCore parallelism is utilized

**Avoid Trainium for:**

- CNN training (CUDA is 3–5× cheaper)
- Modern architectures with LayerNorm, depthwise convolutions, GELU
- Research iteration (compilation time hurts feedback loops)
- Single-model experimentation

### 5.5 Limitations

Our study has several limitations:

1. Single dataset (ChestX-ray14); results may differ for other medical imaging tasks
2. 128×128 resolution; higher resolutions may show different scaling behavior
3. Single seed per configuration; multiple seeds would strengthen statistical claims
4. FP32 only on Trainium; BF16 showed instability but may work with additional tuning

### 5.6 Future Work

We plan the same analysis for Google’s TPUs, which also use XLA compilation and may exhibit similar compatibility constraints.

## 6 Conclusion

We presented the first comprehensive benchmark of AWS Trainium for medical image classification. Our key findings are:

1. **Accuracy parity:** Trainium achieves equivalent accuracy to CUDA for compatible architectures (ConvNeXt-Pico: F1=0.8007 vs 0.8027)
2. **Architecture limits:** Modern CNNs using depthwise convolutions and LayerNorm (ConvNeXt-Nano and larger) fail on Trainium due to hardware constraints
3. **Cost disadvantage:** Trainium is 3–5× more expensive than CUDA for CNN training, contrary to expectations for CNN workloads
4. **Porting complexity:** Four critical XLA-specific code modifications are required, with the validation loop fix particularly non-obvious

For medical imaging practitioners, CUDA remains the recommended platform for CNN-based workflows. Trainium may become more competitive as the Neuron compiler matures, but current limitations make it unsuitable for production medical imaging applications. These findings reflect the Neuron compiler and hardware constraints as of SDK 2.21; improvements in scratchpad management or operator fusion may expand CNN compatibility in future Trainium generations

## Data Availability

All data produced in the present study are available in an AWS S3 bucket, the access method is described in the paper

## Acknowledgments

We thank the NIH Clinical Center for providing the ChestX-ray14 dataset, and Anthropic (Claude.ai) and OpenAI (ChatGPT) for assistance in debugging XLA-specific issues.

## Author Contributions

G.F. conceived the study, designed the experiments, conducted all data analysis, developed the training and evaluation infrastructure, performed the computational experiments, analyzed the results, and wrote the manuscript. All work was conducted independently without collaborators.

## Funding

This research received no external funding and was conducted independently using personal resources.

## Data Availability

The NIH ChestX-ray14 dataset is publicly available at https://nihcc.app.box.com/v/ChestXray-NIHCC.

## Code Availability

Code for reproducing these experiments is available in the AWS S3 bucket `nih-chest-x-rays/trainium` (region: us-east-2).

To list the scripts:

`aws s3 ls s3://nih-chest-x-rays/trainium/scripts/ --no-sign-request --region us-east-2`

To list the output from several runs:

`aws s3 ls s3://nih-chest-x-rays/trainium/results/ --recursive \`

`--region us-east-2 --no-sign-request`

## Competing Interests

The author declares no competing interests. This research was self-funded and not sponsored by AWS, NVIDIA, or any cloud provider.

## Appendices

### A Detailed Per-Disease Results

Table 9 shows the complete per-disease performance metrics for ConvNeXt-Pico trained on Trainium.

**Table 9.** Per-disease performance metrics (ConvNeXt-Pico, Trainium)

| Disease | Prevalence | F1 | ROC-AUC | PR-AUC | Sens. | Spec. |
| --- | --- | --- | --- | --- | --- | --- |
| Hernia | 0.2% | 0.909 | 0.935 | 0.846 | 0.833 | 1.000 |
| Fibrosis | 1.5% | 0.862 | 0.894 | 0.805 | 0.792 | 0.999 |
| Edema | 2.2% | 0.827 | 0.929 | 0.815 | 0.760 | 0.998 |
| Pneumonia | 1.3% | 0.819 | 0.864 | 0.738 | 0.726 | 0.999 |
| Effusion | 13.3% | 0.809 | 0.944 | 0.859 | 0.773 | 0.982 |
| Emphysema | 2.3% | 0.802 | 0.919 | 0.776 | 0.732 | 0.998 |
| Cardiomegaly | 2.5% | 0.801 | 0.911 | 0.776 | 0.722 | 0.998 |
| Consolidation | 4.1% | 0.794 | 0.886 | 0.757 | 0.689 | 0.998 |
| Nodule | 6.1% | 0.778 | 0.862 | 0.741 | 0.669 | 0.997 |
| Mass | 5.1% | 0.778 | 0.880 | 0.757 | 0.670 | 0.997 |
| Atelectasis | 11.2% | 0.770 | 0.909 | 0.811 | 0.708 | 0.985 |
| Pleural Thickening | 2.8% | 0.759 | 0.860 | 0.719 | 0.665 | 0.997 |
| Pneumothorax | 5.3% | 0.751 | 0.907 | 0.754 | 0.651 | 0.996 |
| Infiltration | 19.4% | 0.749 | 0.873 | 0.800 | 0.672 | 0.974 |
| **Macro Average** | — | **0.801** | **0.898** | **0.783** | **0.719** | **0.994** |

## B Project Cost Breakdown

**Table 10.** Total project investment.

| Category | Cost |
| --- | --- |
| AWS compute (g5.2xlarge) | \$50 |
| AWS compute (trn1.2xlarge) | \$100 |
| AWS compute (trn1.32xlarge) | \$2,224 |
| AI assistance (Claude Pro) | \$375 |
| **Total** | **\$2,749** |

## C Statistical Measures

This appendix defines the evaluation metrics used throughout this study. All metrics are computed per-disease and then macro-averaged across the 14 pathology classes.

### C.1 Primary Metrics

#### F1 Score

The harmonic mean of precision and recall, balancing false positives and false negatives:

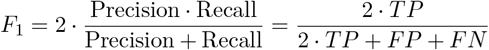

#### ROC-AUC

Area Under the Receiver Operating Characteristic curve, measuring discrimination ability across all classification thresholds. Ranges from 0.5 (random) to 1.0 (perfect).

#### PR-AUC

Area Under the Precision-Recall curve (also called Average Precision). More informative than ROC-AUC for imbalanced datasets where negative cases dominate.

### C.2 Clinical Metrics

#### Sensitivity (Recall, True Positive Rate)

Proportion of actual positives correctly identified:

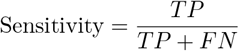

#### Specificity (True Negative Rate)

Proportion of actual negatives correctly identified:

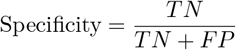

#### Youden’s J Statistic [11]

Summarizes diagnostic effectiveness by combining sensitivity and specificity into a single measure:

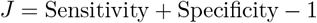

Ranges from 0 (no discriminative ability) to 1 (perfect classification). Often used to select optimal classification thresholds.

### C.3 Derived F-Scores

**F**_***SS***_ (Sensitivity-Specificity F-Score). Harmonic mean of sensitivity and specificity, treating both equally:

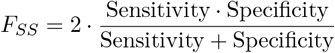

**F**_*β****=2***_ (F-Beta with *β* = 2). Weighted F-score emphasizing recall over precision, appropriate when false negatives are costlier than false positives (common in medical screening):

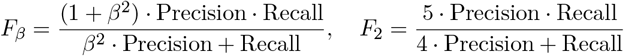

### C.4 Training Stability Metrics

#### Excess Total Variation (ExcessTV)

Measures training curve “wiggliness” beyond net progress, quantifying optimization stability. Given a sequence of validation F1 scores *y*_0_, *y*_1_, …, *y*_*n*− 1_ across epochs:

**Total Variation:**

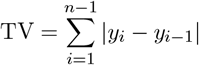

**Net Change:**

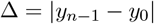

**Excess Total Variation:**

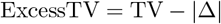

**Interpretation:**

- ExcessTV ≈ 0: Mostly smooth, monotonic improvement
- ExcessTV ≫ 0: Volatile training with many up/down oscillations

The **volatility ratio** TV*/*|Δ| indicates how much “extra distance” the optimization travels relative to its net progress. A ratio of 1.0 represents perfect monotonic convergence; higher values indicate wasted optimization effort due to gradient noise or numerical instability.

This metric is particularly useful for comparing training stability across different precision modes (FP16 vs FP32) or hardware platforms, as lower precision typically introduces more gradient noise and higher ExcessTV.

### C.5 Notation

- TP: True Positives (disease present, correctly predicted)
- TN: True Negatives (disease absent, correctly predicted)
- FP: False Positives (disease absent, incorrectly predicted as present)
- FN: False Negatives (disease present, incorrectly predicted as absent)

All metrics are macro-averaged: computed independently for each of the 14 diseases, then averaged with equal weight regardless of disease prevalence.

## Notes

### Competing Interest Statement

The authors have declared no competing interest.

### Funding Statement

This study did not receive any funding

